# Why (and how) COVID-19 could move us closer to the “health information for all” goal

**DOI:** 10.1101/2020.07.23.20160481

**Authors:** Marco Capocasa, Paolo Anagnostou, Giovanni Destro Bisol

**Author notes:** Corresponding author: Giovanni Destro Bisol, e-mail address.

## Abstract

In this manuscript, we present an analysis of open access (OA) rates of papers concerning COVID-19 and other important human diseases, whose results helped develop an evidence-based scalable strategy aimed at increasing the full and timely access to medical literature. We show that COVID-19 papers are much more openly available (OA rate of 89.5%) than those concerning the four most recent viral outbreaks (Avian influenza, Middle East Respiratory Syndrome, Severe Acute Respiratory Syndrome, Swine influenza; OA rates (from 26.2% to 51.3%) and the ten non COVID-19 disease categories responsible for the highest number of deaths worldwide (OA rates from 44.0% for “Maternal and neonatal disorders” to 58.9% for “Respiratory infections and tuberculosis”). This evidence confronts us with an inevitable question: how can we bridge the gap between OA rates for COVID-19 and other high-impact human diseases? Based on empirical data and projections, we show that it is possible to increase substantially immediate OA to publicly-funded research and complement more demanding initiatives for access to medical literature in developing countries working on the sharing of post-prints at individual, group and multi stakeholder partnership level. However, to make our plan effective in bringing us closer to the “health information for all” goal a more widespread culture of cooperation is fundamental. We argue that the lesson taught by COVID-19 is a unique opportunity to raise awareness among researchers and stakeholders about the importance of open science for human health and to demonstrate that a real change is now possible.

---

Up to July 9 (00:00 GMT), six months after the identification of its etiological agent, the severe acute respiratory syndrome coronavirus 2 (SARS-CoV-2), 11,994,182 cases of Coronavirus disease-19 (COVID-19) and 547,931 related deaths have been reported worldwide.^1,2^ Due to its novelty, the infectivity, pathogenesis and clinical course of COVID-19 remain under scrutiny, and while waiting for a vaccine, a wide range of pharmacological approaches is being tested. In this evolving situation, having readily available new knowledge on COVID-19 may be of great help for all research and intervention activities. Furthermore, combined with effective data sharing, Open Access (OA) to scientific papers could make it possible for more researchers to compare and even check the results of experimental and clinical studies, an essential step towards ensuring scientific integrity and reproducibility.^3,4^ The adoption of OA policies for COVID-19 papers by some major publishers (e.g. Elsevier, the Nature Publishing Group and the American Medical Association) represents a move in this direction. However, the proportion of the peer reviewed literature on COVID-19 that is openly accessible and how this compares to other major diseases is unknown.

To answer these questions, we queried the Web of Science (WoS) database using “COVID-19”, “SARS-CoV-2” and “2019-nCoV” as search terms (accessed on July 9^th^; supplementary Table S1). We retrieved a total of 13,678 items, 9.5 times more than the sum of papers concerning the four most recent viral outbreaks (Avian influenza, Middle East Respiratory Syndrome, Severe Acute Respiratory Syndrome, Swine influenza) in the same length of time (supplementary Table S2), a finding that highlights the very high responsiveness of the scientific community to the new global public health threat. The overall rate of OA COVID-19 papers was 89.5%, whereas those for other viral outbreaks ranged from 26.2% (Severe Acute Respiratory Syndrome) to 51.3% (Swine influenza), with intermediate values for Avian influenza (27.9%) and Middle East Respiratory Syndrome (37.7%).

As we have previously stated concerning the wide openness attitude of human paleo-geneticists^5^, the very high OA rate of COVID-19 papers we observed should not be seen simply as a symbol for the Open Science movement. In fact, it also suggests that the need to open up access to new knowledge regarding high-impact human diseases is finally making headway among researchers and stakeholders.^6^ Although present-day values obviously underestimate the long-term effects of COVID-19, it should be noted that the current number of deaths attributed to the new viral pandemic is 38.7% of the median value for the ten non COVID-19 disease categories responsible for the highest number of deaths worldwide in an equivalent time period (1,414,602; data inferred from 2017 values^7^), while the ratio for projected COVID-19 deaths (1,004,073) up to 01 November 2020 is 43.4% (supplementary Table S3).

With this perspective in mind, we compared OA rates of COVID-19 papers with that of the ten diseases mentioned above. We found substantially lower OA rates for papers concerning non COVID-19 diseases (from 44.0% for “Maternal and neonatal disorders” to 58.9% for “Respiratory infections and tuberculosis”), while only three diseases crossed the 50% threshold (see Fig. 1 and supplementary Table S4). This evidence confronts us with an inevitable question: how can we bridge the gap between OA rates for COVID-19 and other high-impact human diseases? We believe that the following evidence-based strategy, which we developed along a bottom-up-top-down gradient, could be a useful starting point. Essentially, through our proposal, we would like to promote a wider accessibility of post-prints (also referred to as author accepted manuscripts), drafts of articles that have been peer-reviewed and accepted for publication but that have yet to be formatted by the journals.

**Fig. 1.**
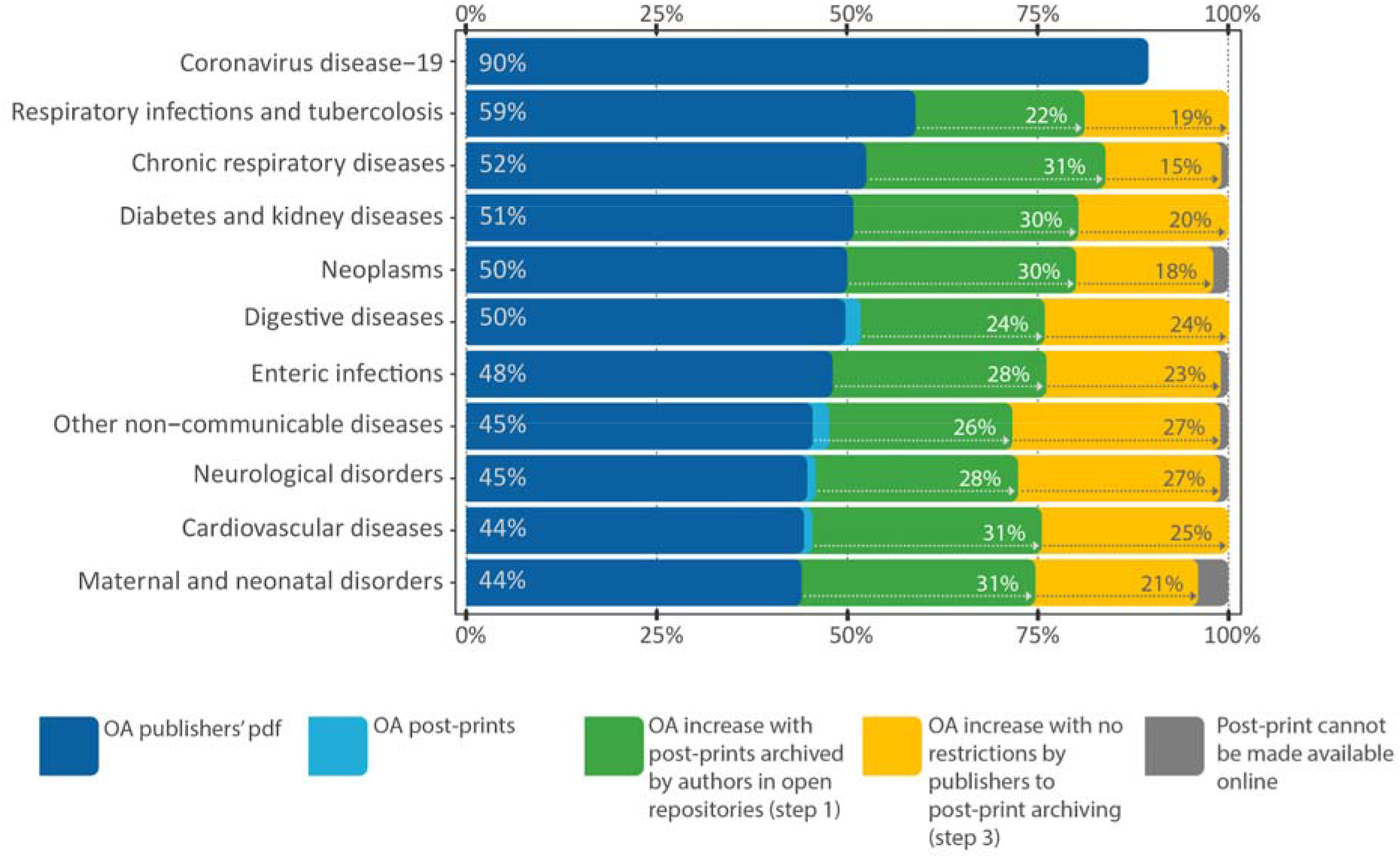
Open Access rates for papers (publishers’ PDFs and post-prints) on COVID-19 and other human diseases. All observed and estimated data are provided in the supplementary material.

Our plan is divided into three scalable steps.

1. At individual level, authors should be more careful about archiving their post-prints in the quickest and most easily accessible way that complies with journals’ rules (http://sherpa.ac.uk/). Interestingly, we randomly selected up to 50 post-prints published in 2020 per each of the ten non COVID-19 diseases considered here (supplementary Table S5). While the overall potential OA increase achievable with post-prints which are immediately shareable according to the journals’ rules is of 29.2%, we could find online only 2.2% of them (six out 273; see Fig. 1 and supplementary Table S6 for disaggregated values). This suggests that we could fill a substantial part of the OA gap with COVID-19 with behavioral changes that are easy to implement and practically at no cost. However, since most journals permit immediate download of post-prints only from personal and departmental web pages, the search engine indexing of the latter should be optimized to make sharing more effective.
2. At group level, any funding agency, academic and research centre, scientific and professional association should promote the open archiving of post-prints by their authors and encourage their dissemination through easily findable online tools.
3. At multi-stakeholder partnership level, academic and research centres should collaborate with scientific associations, patient and professional associations to remove the embargo and other restrictions to online post-print archiving from contracts with publishers, e.g. paying special attention to this point in the negotiation of the “transformative agreements” (https://esac-initiative.org/). This would be another fundamental objective to pursue in light of the evidence that journal rules limited the online archiving of 42.7% of post-prints (210 out of 492) of our dataset (see Fig. 1 and supplementary Table S6 for disaggregated values).

Through the implementation of this strategy, we could complement more demanding initiatives for open access to medical literature. By exploiting the potential of post-prints as a tool for sharing information, we can introduce an alternative and feasible path to achieve immediate open access to publicly-funded research, a need increasingly felt by the scientific community worldwide.^8^ As our observations suggest (see step 1), in most cases the possibility to share post-prints in accordance with the journal rules does not necessarily translate into their open archiving, raising a problem that could be addressed more effectively by individual institutions and their synergic action (see steps 2 and 3) than large-scale initiatives.

With particular regard to developing countries, staff members and students of non-profit institutions (e.g. healthcare centers, professional schools and research institutes) might also use peer-reviewed literature which is not covered by the “Health InterNetwork Access to Research Initiative” (Hinari).^9^ Even researchers and practitioners that are ineligible to join Hinari (e.g. rural and community-based practitioners) and institutions of the least disadvantaged countries that are unable to pay the fees would benefit from more widespread post-print sharing, having a readily available range of information which is significantly wider than that offered by OA journals.^10^ In both cases, increasing post-print availability would reduce the use of the pirated repository of scientific articles.^11^ Finally, the option of sharing of scientific content through post-prints seems to address the concerns recently raised by the European Research Council for the costs that may be created by the adoption of open access policies for countries with more limited financial support for research.^12^

Overall, we believe that these actions could help bring us closer to the “health information for all” goal.^13^ However, such an ambitious aim cannot be achieved without a more widespread culture of cooperation. The lesson taught by COVID-19 offers us a unique opportunity to raise awareness among researchers and all stakeholders about the importance of open science for human health and demonstrate that a real change is now possible.

## Data Availability

All the data used in the study are supplied in the uploaded supplementary material.

## Acknowledgements

This work was supported by the University of Rome “La Sapienza” (Italy), through the project “Setting up a network for Responsible Research and Innovation - RRI-Net” (PI116154C845CA8D).

## Authors’ contributions

MC: initial research idea, help in experimental design, data collection and help in manuscript drafting; PA: data analysis, experimental design and help in manuscript drafting; GDB: experimental design and manuscript writing. All authors revised and approved the manuscript.

